# A comparison of sleep-wake patterns among school-age children and adolescents in Hong Kong before and during the COVID-19 pandemic

**DOI:** 10.1101/2022.06.01.22275778

**Authors:** Steven WH Chau, Samara Hussain, Sandra SM Chan, Oscar WH Wong, Jihui Zhang, Hongliang Feng, Kelly YC Lai, Suk Ling Ma, Suzanne HW So, Kelvin KF Tsoi, Yun Kwok Wing, Se Fong Hung, Patrick WL Leung

## Abstract

**Background:** Lifestyle of children and adolescents have changed extensively during the COVID-19 pandemic due to school suspension and social distancing measures, which can affect their sleep health. Existing studies in the area used convenient samples and focused on the initial months of the pandemic.

**Method:** As part of a territory-wide epidemiological study in Hong Kong, this cross-sectional study recruited primary and secondary school students by stratified random sampling. We investigated the pandemic’s effects on sleep parameters using multivariate regression, adjusting for age, sex, household income, seasonality and presence of mental disorders, and the effects’ moderators and mediators.

**Findings:** Between September 1, 2019 and June 2, 2021, 791 primary and 442 school students were recruited and analysed. After correcting for multiple testing, being assessed during COVID predicted a longer sleep latency in primary and secondary school students in school days (95% CI = 1.0–5.2 minutes, adjusted p-value = 0.010; and 95% CI= 3.9–13.0 minutes, adjusted p-value =0.004, respectively) and non-school days (95% CI = 1.7–7.2 minutes, adjusted p-value = 0.005; 95% CI = 3.4–13.7 minutes, adjusted p-value = 0.014, respectively). Low household income was a moderator for later bedtime (adjusted p-value = 0.032) and later sleep onset (adjusted p-value = 0.043) during non-school days among secondary school students. Sex and digital leisure time were not moderator and mediator of the pandemic’s effect on sleep parameters, respectively.

**Interpretation:** Changes associated with the COVID-19 pandemic have a widespread and enduring effect on sleep health of school-aged students in Hong Kong. Household income play a role in adolescents’ sleep health’s resilience against these changes, and anti-epidemic measures effects on the health gap of the youth should be considered.

**Funding:** Government of the Hong Kong Special Administrative Region, Food and Health Bureau, Health and Medical Research Fund (Ref. No.: MHS-P1(Part 1)-CUHK).

---

By now, the world has lived with the COVID-19 pandemic for over two years. While children and adolescents may not be worst hit by direct COVID-19 related health problems due to the generally milder disease severity among this population if infected (1), they suffer from indirect effects on their lifestyle due to widespread school suspensions, home confinement and reduced accessibility to healthcare and social resources. In March 2021, UNICEF estimated that 168 million children could not attend school for one year (2). A wide range of health problems have been rising among children and adolescents globally due to such disruption (3,4). Sleep health, an essential aspect of health, is sensitive to lifestyle changes. The sleep-wake patterns of young individuals are highly affected by their daytime activity routine, school schedules and the lifestyles of their parents (5). Disruptions of sleep and circadian rhythms in the younger population have significant physical and mental health implications (6,7). It is, therefore, important to examine changes in sleep and circadian parameters during the COVID-19 pandemic and to identify biopsychosocial risk factors for a more precise formulation of preventive public health measures. A meta-analysis of nine studies estimated that up to 54% of school-aged youths suffer from sleep problems during the early stage of the pandemic (8). However, existing studies were limited by methodological concerns such as the use of convenient sampling methods and retrospective recalls, making them prone to biases. Second, these studies covered the first months of the pandemic only, and an updated investigation would be timely to reflect the ‘new normal’ of living with the pandemic as it lingered on. Third, these studies did not set out to identify factors moderating the effect of the COVID pandemic on young people’s sleep health.

Biological sex is one of the factors that potentially moderate the effect of pandemic associated changes on young people, since it is known that insomnia is more common among females than males since puberty (9). Another potential moderating factor is household income. Low household income is a predictor of poorer health outcomes for young people (10). During the pandemic, socioeconomic status is repeatedly shown to be a key determinant for various infection risks and infection outcomes worldwide, including in Hong Kong (11). For young people specifically, school closure and the social restrictions would likely cause more activity limitations for those with fewer financial resources (12).

Hong Kong had been spared of massive causalities from COVID-19 until the recent 2022 surge due to the Omicron variant, hence avoiding a hard lockdown. However, prolonged and stringent social distancing measures and school suspensions have been in place over the majority of the time between early 2020 and mid-2021 (Figure 1). School-age children and adolescents were directly affected by some of these measures, especially the year-long school suspension and the adoption of online home learning. There were concerns that these changes would disrupt the lifestyles of children and adolescents, and consequently threaten their health, including sleep health. These changes may also lead to an increase in digital leisure time, exacerbating sleep problems (13).

**Figure 1:**
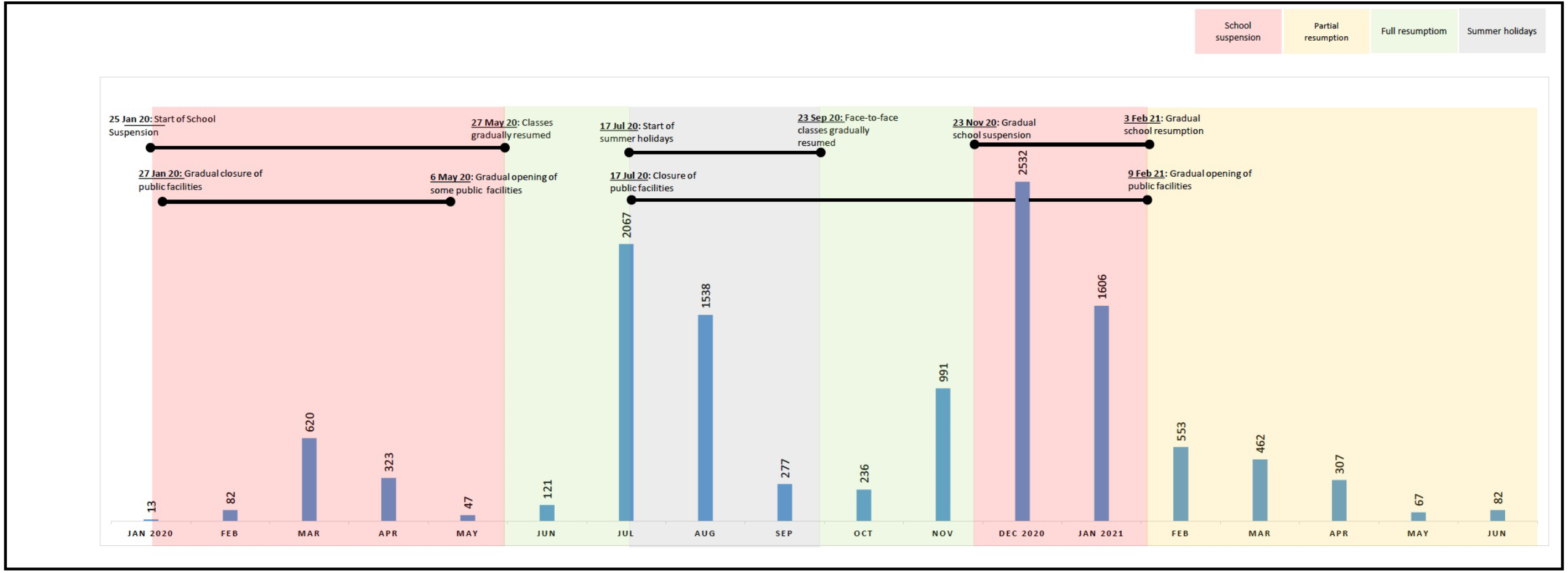
Timeline of anti-pandemic measures from January 2020 to June 2021 in Hong Kong. Bar charts and numbers represent the total number of confirmed COVID-19 cases of the month.

The “*Let’s Work Heart*” project is a territory-wide epidemiological study of mental, sleep and behavioural health among primary and secondary school students in Hong Kong. The study began in 2019 (i.e. before the pandemic started) and is still ongoing. The timeframe of the study thus provides a natural opportunity to compare the sleep-wake patterns in a representative sample of school-age children and adolescents of Hong Kong between the pre-COVID and COVID periods. In this sub-study, we asked three questions: (1) are there differences in sleep-wake patterns among school students during COVID times as compared to the pre-COVID period; (2) are the differences in these patterns, if any, different between primary and secondary school students; (3) do sociodemographics moderate the differences in sleep parameters before and during COVID times; and (4) does change in the amount of digital leisure time mediate the differences in sleep parameters before and during COVID times, if any.

## Method

This is a cross-sectional study.

### Participants

The inclusion criteria were students aged 6-17 years from any mainstream primary school and secondary school in Hong Kong, where school enrollment in this age group is universal (14). No exclusion criteria were applied. The study adopted a two-stage random sampling method. Schools were first randomly selected using stratified sampling. Within each participating school, every 1 out of 7 students was then randomly selected. Written parental consent was obtained from all student participants. The study was approved by the Joint Chinese University of Hong Kong – New Territories East Cluster Clinical Research Ethics Committee (Ref. no: 2018.497).

### Measures

We record demographic and socioeconomic data of participants’ households, including household size and household income. Students’ electronic media use was assessed by the Media Activity Form (MAF) (16). For primary school student participants, one of their parents (or the primary caregiver) was interviewed by trained research staff for the presence of lifetime sleep disorders using the modified version of the structured diagnostic interview for sleep patterns and disorders (DISP)(15). The DISP inquires into the sleep-wake schedules of the student participant during school days and non-school days in the past month. The sleep parameters we investigated were bedtime, sleep latency, sleep onset time, wake up time, sleep duration and sleep midpoint of school days and non-school days, and social jetlag (i.e., the difference in sleep midpoint between school days and non-school days). They were also interviewed for the presence of psychiatric disorders as defined by the 5^th^ version of the Diagnostic and Statistical Manual (DSM-5) in the past 12 months using the Diagnostic Interview Schedule for Children, 5^th^ version (DISC-5). For secondary school participants, interviews for sleep patterns were conducted with the students themselves, and a multi-informant approach was used for diagnostic interviews for mental disorders (16). All interviews took place at the participant’s school in the pre-COVID phase, while video-conferencing was adopted during the COVID phase.

### Statistical analyses

Data from primary and secondary school students were analysed separately due to significant differences in school-related factors. Subjects with missing sleep parameters are excluded from the analysis. Other missing variables are imputed using median values of the whole data pool. We performed univariate analyses for differences in sleep parameters between participants assessed in the pre-COVID and COVID period using independent t-tests to select sleep parameters of interest for multivariate analyses (criteria set as *p*<0.1). We used 1st March 2020 as a cutoff for the pre-COVID and COVID periods, as the first school suspension in the city started at the end of January 2020 (No interviews were conducted between 6 February 2o2o to 4 March 2o2o)(Supplementary table 1). We then used multivariate general linear models to compute the adjusted regression coefficient of the assessment period as a predictor of the differences in the selected sleep parameters of interest, with potential confounders of age, sex, seasonality (summer/winter/others), income status (low income/middle-to-high income) and presence of any mental disorder being controlled. We also report the more conservative adjusted p-value using the Holm-Sidak method of multiple hypothesis-testing corrections. To examine the moderating effects of sex and household income status, the multivariate general linear models were tested again with interaction terms added. To examine the mediating effect of the amount of digital leisure time, we followed the method described by Baron and Kenny (17). The participants’ amount of digital leisure time was retrieved from the MAF (excluding educational use of digital media). Since the digital leisure time measure was highly skewed, we performed log-transformation on this data before we proceeded with further analysis.

## Results

Over the period from 1^st^ September 2019 to 2^nd^ June 2021 (i.e., over two academic years), we recruited and assessed 793 primary school students and 444 secondary school students. In the specified period, the overall school-level response rate was 75%, and the student-level response rate was 79.5% (recruitment flowcharts in Figures 2a and 2b). Please see Supplementary table 1 for month-by-month recruitment number.

**Figure 2:**
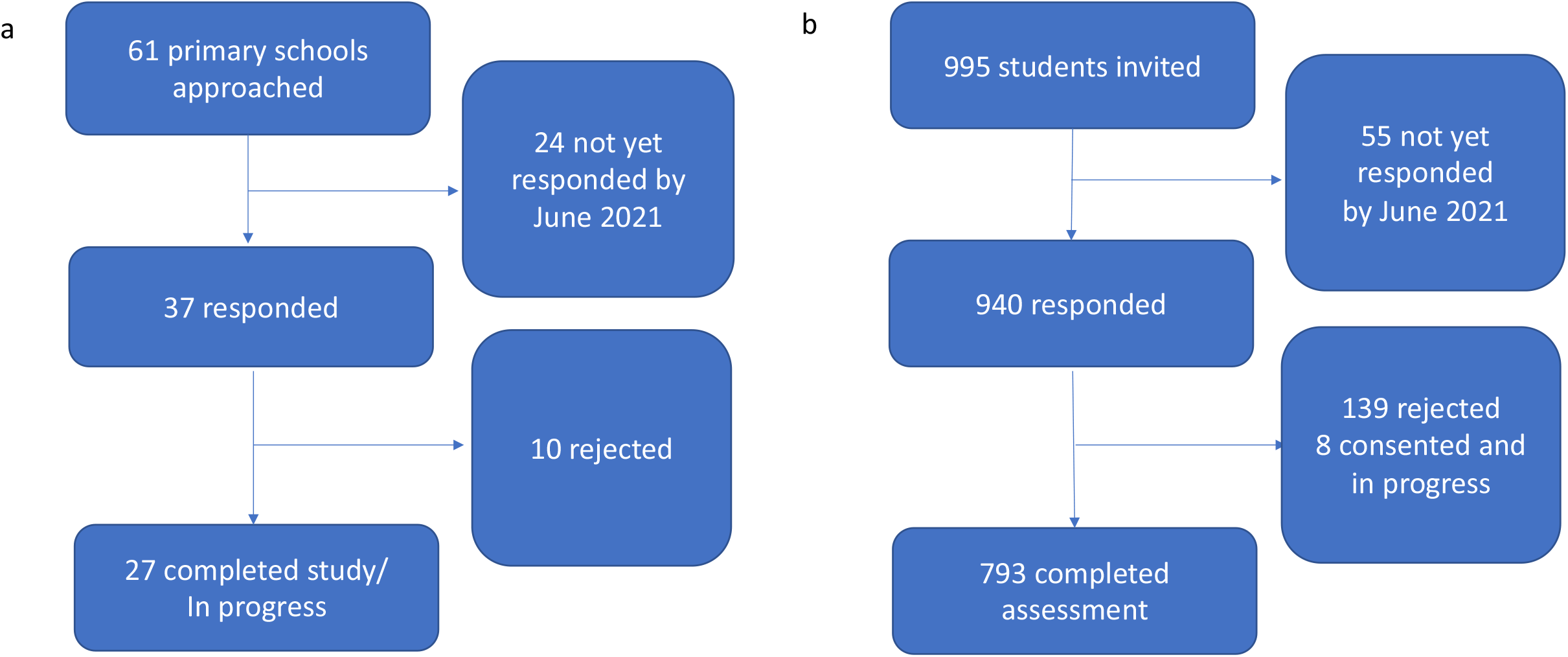
Recruitment flowchart of primary school student subject. a. School-level recruitment; b. Student-level recruitment.

**Figure 3:**
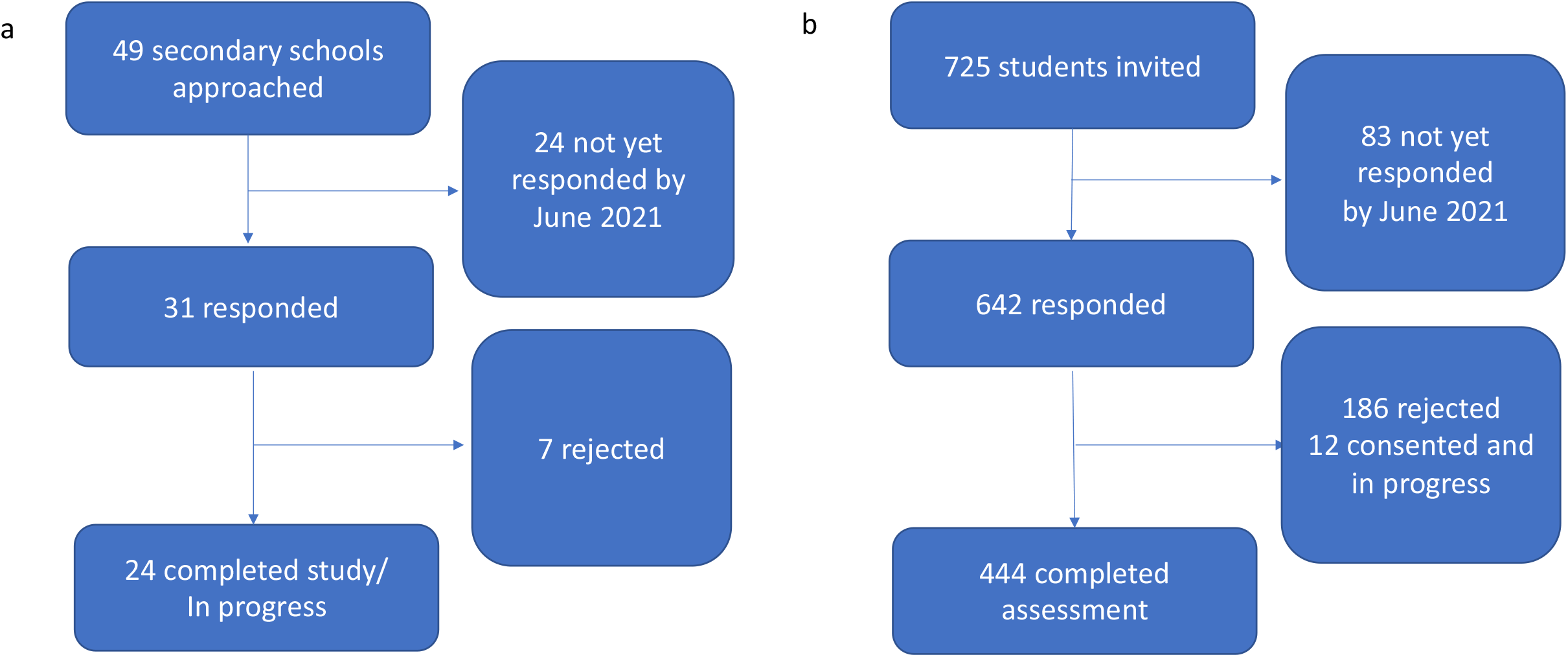
Recruitment flowchart of secondary school student subject. a. School-level recruitment; b. Student-level recruitment.

### Sample characteristics

The mean age was 8.7 years (range 6-13) among the primary school students, and 49.5% were female. Assessment of 247 was performed pre-COVID (before 1^st^ March 2020), and 546 since COVID. Two were excluded for invalid data (one was evaluated in the pre-COVID period, and another in the COVID period), and hence the total analysed sample size was 791. The pre-COVID and COVID period samples were comparable in age, sex, school type, and proportion of low-income households (defined as falling below 50% of the median Hong Kong household income of households of the same size, using data from the 4^th^ Quarter of 2020) (C&SD, HKSAR), but not for the season at the time of assessment and the presence of any mental disorder as defined by the DSM-5 (Table 1a).

**Table 1a:** Comparison of primary school student characteristics pre- and during COVID. Independent t-test were used for all item except for digital leisure time, of which Mann-Whitney U test was used.

|  | Pre-COVID (n=246) | COVID (n=545) | p-value |
| --- | --- | --- | --- |
| Age, years, mean (SD) | 8.5 (1.7) | 8.8 (1.8) | 0.05 |
| Female, n (%) | 120 (48%) | 272 (50%) | 0.75 |
| Government-subsidised school, n (%) | 246 (100%) | 545 (100%) | 1 |
| Low-income household*, n (%) | 71 (28.9%) | 136 (25.0%) | 0.25 |
| Season at the time of assessment: |  |  | **<0.001** |
| Spring (Mar-Apr), n (%) | 0 (0%) | 79 (14.5%) |  |
| Summer (May-Sept), n (%) | 34 (13.8%) | 158 (29.0%) |  |
| Autumn (Oct-Nov), n (%) | 31 (12.6%) | 110 (20.2%) |  |
| Winter (Dec-Feb), n (%) | 181 (73.6%) | 198 (36.3%) |  |
| Any mental disorder, n (%) | 84 (34.1%) | 219 (40.1%) | **0.027** |
| Digital leisure time, hours per week, median | 15.5 | 24.2 | **<0.001** |

The mean age was 14.1 years (range 11-17) among the secondary school students, and 43.7% were female. We assessed 230 of them pre-COVID, and 214 were evaluated during COVID. Two were excluded for invalid data (both were assessed during COVID). Thus, the number of analysed samples was 442. The pre-COVID and COVID period samples matched in school type and frequency of mental disorder, but not age, sex, the proportion of low-income households and season at the assessment time (see Table 1b). Please see supplementary Figure. 2 for summary of number of missing data of all variables involved in the analysis

**Table 1b:** Comparison of secondary school student characteristics pre- and during COVID. Independent t-test were used for all item except for digital leisure time, of which Mann-Whitney U test was used.

|  | Pre-COVID (n=230) | COVID (n=212) | p-value |
| --- | --- | --- | --- |
| Age, years, mean (SD) | 13.9 (1.8) | 14.3 (1.7) | **0.045** |
| Female, n (%) | 111 (48%) | 82 (39%) | **0.042** |
| Government-subsidised school, n (%) | 230 (100%) | 212 (100%) | 1 |
| Low-income household*, n (%) | 58 (25.2%) | 80(37.7%) | 0.005 |
| Season at the time of assessment: |  |  | **<0.001** |
| Spring (Mar-Apr), n (%) | 0 (0%) | 40 (18.9%) |  |
| Summer (May-Sept), n (%) | 64 (27.8%) | 44 (20.8%) |  |
| Autumn (Oct-Nov), n (%) | 88 (41.5%) | 19 (9.0%) |  |
| Winter (Dec-Feb), n (%) | 78 (33.9%) | 109 (36.3%) |  |
| Any mental disorder, n (%) | 83 (36.1%) | 73 (51.4%) | **0.72** |
| Digital leisure time, hours per week, median | 64.0 | 71.4 | **0.043** |

### Comparison of sleep parameters before and during COVID

Univariate analysis showed that in comparison to those assessed pre-COVID, primary students assessed in COVID times had significantly longer sleep latency on school days (*p* <0.001) and non-school days (p < 0.001), and longer sleep duration on non-school days (*p* = 0.026) (Table 2a).

**Table 2a:** Comparison of sleep parameters among primary school students pre- and during COVID. Independent t-test was used for all items.

|  | Pre-COVID (n=246)<br>*M (SD)* | COVID (n=545)<br>*M (SD)* | *p*-value |
| --- | --- | --- | --- |
| School days |  |  |  |
| Bedtime, hh:mm | 22:08 (±45 mins) | 22:09 (±51 mins) | 0.84 |
| Sleep onset time, hh:mm | 22:22 (±47 mins) | 22:27 (±51 mins) | 0.25 |
| Sleep midpoint time, hh:mm | 2:42 (±29 mins) | 2:42 (±37 mins) | 0.94 |
| Wake up time, hh:mm | 7:01 (±37 mins) | 6:57 (±25 mins) | 0.13 |
| Sleep latency, mm:ss | 0:13 (±12 mins) | 0:17 (±13 mins) | **<0.001** |
| Sleep duration, hh:mm | 8:39 (±48 mins) | 8:30 (±50 mins) | **0.03** |
| Non-school days |  |  |  |
| Bedtime, hh:mm | 22:56 (±53 mins) | 22:55 (±61 mins) | 0.91 |
| Sleep onset time, hh:mm | 23:12 (±56 mins) | 23:17 (±62 mins) | 0.31 |
| Sleep midpoint time, hh:mm | 3:59 (±56 mins) | 4:06 (±60 mins) | 0.17 |
| Wake up time, hh:mm | 8:47 (±70 mins) | 8:55 (±74 mins) | 0.17 |
| Sleep latency, mm:ss | 0:16 (±15 mins) | 0:21 (±17 mins) | **<0.001** |
| Sleep duration, hh:mm | 9:34 (±59 mins) | 9:37 (±65 mins) | 0.53 |
| Social jetlag, hh:mm | 1:17 (±52 mins) | 1:23 (±48 mins) | 0.11 |

**Table 2b:** Comparison of sleep parameters among secondary school students pre- and during COVID. Independent t-test was used for all items.

|  | Pre-COVID (n=230)<br>*Mean (SD)* | COVID (n=212)<br>*Mean (SD)* | *p*-value |
| --- | --- | --- | --- |
| School days |  |  |  |
| Bedtime, hh:mm | 23:13 (±72mins) | 23:24 (±82 mins) | 0.13 |
| Sleep onset time, hh:mm | 23:27 (±73 mins) | 23:46 (±91 mins) | **0.016** |
| Sleep midpoint time, hh:mm | 3:03 (±43 mins) | 3:15 (±55 mins) | **0.015** |
| Wake up time, hh:mm | 6:39 (±30 mins) | 6:43 (±46 mins) | 0.28 |
| Sleep latency, hh:mm | 0:14 (±12 mins) | 0:22 (±31 mins) | **<0.001** |
| Sleep duration, hh:mm | 7:11 (±70 mins) | 6:56 (±94 mins) | 0.061 |
| Non-school days |  |  |  |
| Bedtime, hh:mm | 00:01 (±84 mins) | 00:28 (±85 mins) | **0.006** |
| Sleep onset time, hh:mm | 00:16 (±85 mins) | 00:51 (±125 mins) | **<0.001** |
| Sleep midpoint time, hh:mm | 4:41 (±81 mins) | 5:05 (±100 mins) | **0.005** |
| Wake up time, hh:mm | 9:20 (±105 mins) | 9:43 (±112 mins) | **0.030** |
| Sleep latency, hh:mm | 00:14 (±13 mins) | 00:22 (±35 mins) | **0.002** |
| Sleep duration, hh:mm | 9:03 (±102 mins) | 8:51 (±105 mins) | 0.23 |
| Social jetlag, hh:mm | 1:37 (±104 mins) | 1:50 (±137 mins) | **0.049** |

Using a multivariate linear regression, assessment period (i.e., whether one is assessed in the pre-COVID or during COVID) remained a significant predictor of longer sleep latency on school days (*B* = 3.1 minutes, 95% CI = 1.0–5.2, *p* = 0.005) and non-school days (*B* = 4.4 minutes, 95% CI = 1.7–7.2, *p* = 0.002), but not for sleep duration (*B* = 5.9 minutes, 95% CI = -13.6–1.9, *p* = 0.13). After applying Holm-Sidak correction, assessment period remained a significant predictor of longer sleep latency during both school days and non-school days (adjusted *p*-values = 0.010 and 0. 005 respectively).

Univariate analysis showed that secondary school students assessed during the pandemic had longer self-reported sleep latency (*p* < 0.001), later sleep onset time (*p*= 0.016) and a later sleep midpoint (*p* = 0.015) on school days, compared to their peers who were assessed pre-COVID. On non-school days, students assessed in the COVID-period reported a later bedtime (*p* = 0.006), longer sleep latency (*p* = 0.002), later sleep onset time (*p* < 0.001), later wake up time (*p* = 0.030) and a later sleep midpoint (*p* = 0.005) than their peers who were assessed pre-COVID. Social jetlag was longer among students assessed in the COVID era (p = 0.049) (Table 3).

Using a multivariate regression model, assessment period was a significant predictor of later bedtime on non-school days (*B*= 21.3 minutes, 95% CI = 2.9–39.6, *p* = 0.023), longer sleep latency on both school days (*B* = 8.5 minutes, 95% CI= 3.9–13.0, *p* <0.001) and non-school days (*B* = 8.6 minutes, 95% CI = 3.4–13.7, *p* = 0.001), later sleep onset on both school days (*B* = 17.0 minutes, 95% CI = 12.8–21.0, *p* = 0.025) and non-school days (*B* = 29.8 minutes, 95% CI = 10.1–49.6, *p* = 0.0031), and a later sleep midpoint on both school days (*B* = 10.1 minutes, 95% CI = 1.8–19.9, *p* = 0.019) and on non-school days(*B* = 19.5 minutes, 95% CI = 2.4–36.6, *p* = 0.025), and shorter sleep duration during school days (*B* = 19.5 minutes, 95% CI = 2.4–36.6, *p* = 0.019). After applying Holm-Sidak correction, assessment period remains a significant predictor of longer sleep latency during both school days (adjusted p-value =0.004) and non-school days (adjusted *p*-value =0.014) and later sleep onset during non-school days (adjusted *p*-value =0.025).

### Moderators of differences in sleep parameters

For both primary and secondary school groups, multivariate regression models containing assessment period x sex interaction term did not find participants’ sex to significantly moderate the COVID era’s predictive effect for any sleep parameters (*p* > 0.05).

Among primary school students, multivariate regression models containing assessment period x household income status interaction term showed that low household income is not a significant moderator of the effect of assessment period as a predictive factor of the sleep parameters of interest (*p-values* > 0.05).

Among secondary school students, the same multivariate regression models showed that being assessed during the pandemic was significantly more predictive of a later bedtime (*B* = 56.7 minutes, 95% CI = 18.2–95.3, *p* = 0.004), sleep onset (*B* = 56.7 minutes, 95% CI = 15.3–98.2, *p* = 0.0071) and sleep mid-point (*B* = 37.2 minutes, 95% CI = 1.1–73.2, *p* =0.044) during non-school days among students from low-income households, compared to their peers from middle-to-high-income families. After applying the Holm-Sidak correction, low household income remained a significant moderator for later bedtime (adjusted p-value = 0.032) and later sleep onset (adjusted p-value = 0.043) on non-school days among secondary school students.

### Digital leisure time as a mediator of differences in sleep parameters

The median weekly digital leisure time of the pre-COVID and COVID period sample of primary school students are 15.5 hours and 24.2 hours, respectively. The median digital leisure time of the pre-COVID and COVID period sample of secondary school students are 64.0 hours and 71.4 hours, respectively. While univariate Mann-Witney U test showed that the median of weekly digital leisure time was significantly different pre- and during COVID for the primary (p<0.001) and secondary school students (p=0.043), using a multivariate regression model containing age, sex, seasonality, and household income as covariates, the assessment period did not predict digital leisure time (log-transformed) changes for primary and secondary school students.

## Discussion

To the best of our knowledge, this is the first study comparing sleep-wake patterns before and after the COVID outbreak among children and adolescents using school-based random sampling.

### Sleep-wake patterns altered during COVID-era for students in both primary and secondary school

Both primary and secondary school students took longer to fall asleep after the COVID outbreak. Such differences remained robust after adjusting for confounders. In our city, characterised by a low infection rate but stringent anti-pandemic measures, pandemic related stressors and lifestyle changes have a widespread effect on the sleep health of young people across ages.

The sleep-wake pattern alteration among secondary school students was more pervasive than the primary school students. With later bedtime, longer sleep latency, later sleep onset and later sleep midpoints, this suggests a pattern of a rearward shift of sleep period during both school days and non-school days, even though only an increase in sleep latency survived correction for multiple comparisons. Our findings extended the current literature that children and adolescents not only had delayed sleep in the first months of the pandemic (18)(19) by demonstrating enduring changes in sleep-wake patterns over the first 15 months of COVID. It is not surprising that secondary school students are affected more than primary school students, as adolescents are more prone to insomnia and sleep phase delays than younger children (6)(9). Under COVID, young people in Hong Kong experienced profound lifestyle changes: school attendance was suspended, online learning via video-conferencing platforms became the “new normal”, and activities outside the home were restricted due to social distancing measures and the closing down of venues. As physical activities and light exposure are key inputs of our circadian clock, these changes can disturb the circadian rhythm of young people (20). Stress from fear of infection, social isolation and uncertainty are also possible factors that contribute to the increase in sleep difficulties.

### Secondary school students from low-income households had more delayed sleep on non-school days during COVID

Our moderation analysis suggested that the sleep-wake pattern of boys and girls studying in both primary and secondary school were equally altered during the COVID pandemic. Analysis of the moderating effect of household income revealed that secondary school students from low-income households have a consistent pattern of more delayed sleep on non-school days during the COVID period than their peers from higher-income households, but not on school days. Sleep-wake pattern during the non-school day is a better indicator of one’s ‘biological clock’ since one is less confined by the ‘social clock’. However, the increase in sleep latency during both school days and non-school days was not moderated by household income. Therefore, our result suggests that secondary school students from low-income households have a more pronounced backward circadian rhythm shift than their wealthier peers without an increase in difficulty in sleep initiation. We speculate that this shift in circadian rhythm could be understood in our city’s context of social inequality, where low-income families were more deprived of social and physical activities during the pandemic. Like many urban areas worldwide, children and adolescents from low-income households in Hong Kong rely more on school-based support for activity opportunities. Data from the US showed that children from low-income families who cannot attend school physically during COVID suffer from more marked mental health impairment (21). Besides, youths from low-income households were more entrapped at home than their peers from higher-income families due to the closing of government-administered leisure venues over an extended period. This could have widened the gap in accessibility to activities between the rich and the poor, which can be translated into their differences in the level of physical activities and light exposure, two key zeitgebers of circadian rhythm (22).

Socioeconomic status has always been a significant determinant of health in children and adolescent populations, and it undermines one’s resilience against adverse outcomes in times of stress and crisis (23). In line with our finding on the effect of socioeconomic status on sleep health during COVID, a local cross-sectional online survey found that a low-income status and several other socioeconomic adversities increased the risk of psychosocial problems in children during the COVID pandemic (24). Society might bear the toll of these unintended mental health consequences of widening the health gap between rich and poor young people in the COVID era.

### The amount of digital leisure time did not mediate the effect of COVID-era on the sleep-wake patterns of children and adolescents

Contrary to our expectations, we did not see a significant increase in digital leisure time after the COVID outbreak. As a recent study showed that the rise in use of digital media during the initial lockdown period almost returned to the pre-COVID level afterwards (25), it is possible that during the 15 months of our recruitment during the COVID period, digital leisure time of our sample increased initially and subsequently decreased.

### Sleep-wake pattern disruption and its health effect on children and adolescents

Sleep and circadian health are closely related to children’s and adolescents’ physical and mental health. Delayed sleep preference is associated with a wide range of physical, psychological and behavioural problems among young people, such as depression, substance use (26), and adverse endocrine and physical profiles (27,28). Insomnia, on the other hand, is independently associated with an increased risk of a wide range of emotional and behavioural problems, such as depression, anxiety, daytime sleepiness and suicidality among adolescents (7,29). Evidence from prospective cohort studies suggests that poor sleep and circadian health could be the precedence for at least some of these adverse outcomes (26,30). We should thus not overlook the sleep and circadian health of our young people during the pandemic, particularly when formulating measures that would significantly impact their lifestyle and activities. Even in desperate circumstances when infection control is of overriding priority, the government must consider mitigating measures such as extra allowance for outdoor activities(31) and ensure fairness in the opportunity to access activities, especially for the underprivileged.

### Strengths and Limitations

The key strength of this study is the use of a territory-wide, random sampling method. Together with a high response rate, we have captured a representative sample of school-aged children and adolescents in Hong Kong. Furthermore, we kept our sampling methodology consistent before and during COVID, making our data from the two periods comparable. The study has several limitations: First, this study was cross-sectional by nature and not longitudinal. Second, the sleep parameters were derived only from self-report or parent-report, without objective measures such as actigraphy. Third, our data did not capture daytime activities, limiting our interpretation of mediating factors of changes.

## Conclusion

Our results highlighted demonstrable differences in the sleep patterns of children and adolescents of Hong Kong before and during COVID, despite the low COVID infection rate. These changes can, in turn, lead to adverse health consequences. The moderating effect of household income on circadian disruption among adolescents is a source of concern. It should alert us to the widening health gap between rich and poor youth under the pressure of COVID. Proactive measures to alleviate the health impact of the younger population during COVID should be part of a holistic pandemic response plan.

## Supporting information

STROBE checklist

## Data Availability

All data produced in the present study are available upon reasonable request to the authors

