## Supplementary Table 1 for "A comparison of sleep-wake patterns among school-age children and adolescents in Hong Kong before and during the COVID-19 pandemic"

Supplementary Table 1: Month-by-month recruitment number

| Month | Year | SEC | PRI |
|---|---|---|---|
| **9** | 2019 | 64 | 34 |
| **10** | 2019 | 26 | 3 |
| **11** | 2019 | 62 | 28 |
| **12** | 2019 | 42 | 63 |
| **1** | 2020 | 36 | 118 |
| **2** | 2020 | 0 | 1 |
| **3** | 2020 | 1 | 22 |
| **4** | 2020 | 6 | 11 |
| **5** | 2020 | 9 | 5 |
| **6** | 2020 | 14 | 8 |
| **7** | 2020 | 8 | 32 |
| **8** | 2020 | 3 | 35 |
| **9** | 2020 | 0 | 35 |
| **10** | 2020 | 0 | 26 |
| **11** | 2020 | 11 | 84 |
| **12** | 2020 | 42 | 104 |
| **1** | 2021 | 69 | 63 |
| **2** | 2021 | 7 | 31 |
| **3** | 2021 | 16 | 34 |
| **4** | 2021 | 18 | 12 |
| **5** | 2021 | 8 | 43 |
| **6** | 2021 | 2 | 1 |
| | **Total** | **444** | **793** |
